# Association Between COVID-19 During Pregnancy and Preterm Birth by Trimester of Infection: A Retrospective Cohort Study Using Longitudinal Social Media Data

**DOI:** 10.1101/2023.11.17.23298696

**Authors:** Ari Z. Klein, Shriya Kunatharaju, Su Golder, Lisa D. Levine, Jane C. Figueiredo, Graciela Gonzalez-Hernandez

**Author notes:** Corresponding authors: Ari Z. Klein, University of Pennsylvania, 405 Blockley Hall, 423 Guardian Dr., Philadelphia, PA 19104;, Graciela Gonzalez-Hernandez, Cedars-Sinai Medical Center, Pacific Design Center, Ste. G549F, 700 N. San Vicente Blvd., West Hollywood, CA 90069.

## Abstract

**Background:** Preterm birth, defined as birth at <37 weeks of gestation, is the leading cause of neonatal death globally and, together with low birthweight, the second leading cause of infant mortality in the United States. There is mounting evidence that COVID-19 infection during pregnancy is associated with an increased risk of preterm birth; however, data remain limited by trimester of infection. The ability to study COVID-19 infection during the earlier stages of pregnancy has been limited by available sources of data. The objective of this study was to use self-reports in large-scale, longitudinal social media data to assess the association between trimester of COVID-19 infection and preterm birth.

**Methods:** In this retrospective cohort study, we used natural language processing and machine learning, followed by manual validation, to identify pregnant Twitter users and to search their longitudinal collection of publicly available tweets for reports of COVID-19 infection during pregnancy and, subsequently, a preterm birth or term birth (i.e., a gestational age ≥37 weeks) outcome. Among the users who reported their pregnancy on Twitter, we also identified a 1:1 age-matched control group, consisting of users with a due date prior to January 1, 2020—that is, without COVID-19 infection during pregnancy. We calculated the odds ratios (ORs) with 95% confidence intervals (CIs) to compare the overall rates of preterm birth for pregnancies with and without COVID-19 infection and by timing of infection: first trimester (weeks 1-13), second trimester (weeks 14-27), or third trimester (weeks 28-36).

**Results:** Through August 2022, we identified 298 Twitter users who reported COVID-19 infection during pregnancy, a preterm birth or term birth outcome, and maternal age: 94 (31.5%) with first-trimester infection, 110 (36.9%) second-trimester infection, and 95 (31.9%) third-trimester infection. In total, 26 (8.8%) of these 298 users reported preterm birth: 8 (8.5%) were infected during the first trimester, 7 (6.4%) were infected during the second trimester, and 12 (12.6%) were infected during the third trimester. In the 1:1 age-matched control group, 13 (4.4%) of the 298 users reported preterm birth. Overall, the risk of preterm birth was significantly higher for pregnancies with COVID-19 infection compared to those without (OR 2.1, 95% CI 1.06-4.16). In particular, the risk of preterm birth was significantly higher for pregnancies with COVID-19 infection during the third trimester (OR 3.17, CI 1.39-7.21).

**Conclusion:** The results of our study suggest that COVID-19 infection particularly during the third trimester is associated with an increased risk of preterm birth.

## Introduction

Preterm birth, defined as birth at <37 weeks of gestation, is the leading cause of neonatal death globally^1^ and, together with low birthweight, the second leading cause of infant mortality in the United States.^2^ According to recent systematic reviews and meta-analyses,^3–13^ there is mounting evidence that COVID-19 infection during pregnancy is associated with an increased risk of preterm birth; however, data remain limited on the risk of preterm birth by trimester of infection. As a limitation of their review, Allotey et al.^5^ wrote: “Not many studies reported outcomes by trimester for symptom onset.” Jeong and Kim^13^ also noted that they “did not analyze infections according to pregnancy trimesters.” Among studies that did report the trimester of infection, most of the infections were limited to the third trimester. Sturrock et al.^12^ “were unable to examine the impact by trimester of maternal SARS-CoV-2 infection due to a paucity of studies examining offspring of first or second trimester infection.” Likewise, Smith et al.^10^ wrote: “There were relatively few instances of SARS-CoV-2 infection identified during the first trimester; the majority of cases were identified during the third trimester.” As Marchand et al.^4^ further pointed out, this lack of data affects the generalizability of the available evidence on the association between COVID-19 infection during pregnancy and preterm birth, arguing that “most of the included pregnant women were in the third trimester, so the results of this meta-analysis cannot be generalized to pregnant women in the first and second trimesters.” The ability to study COVID-19 infection during the first and second trimesters has been limited by available sources of data.

Most studies of COVID-19 infection during pregnancy have been limited to the third trimester because their data were collected in the hospital around the time of delivery. Allotey et al.^5^ noted that studies “primarily reported on pregnant women who required visits to hospital, including for childbirth.” Hospital-based data not only limits the timing of COVID-19 infection primarily to the third trimester, but also fails to account for potential exposure during the earlier stages of pregnancy in the comparator group of those admitted to the hospital without infection. As Allotey et al.^5^ have suggested, “collection of maternal data by trimester of exposure, including the periconception period, is required.” To complement pregnancy registries,^14^ our prior work^15^ demonstrated that Twitter can be used as a source of self-report data to assess the association between medication exposure during pregnancy and birth defects, including by trimester of exposure. Our ability to retrospectively collect users’ tweets enables us to observe reports of exposures in the early stages of pregnancy—prior to prenatal care^16^ and even knowledge of being pregnant—while reducing recall bias. While Twitter data has been used for a wide range of studies related to COVID-19,^17^ it has not been used to study COVID-19 infection during pregnancy. The objective of this study was to use large-scale, longitudinal social media data to assess the association between COVID-19 infection during pregnancy—specifically, by trimester of infection—and preterm birth.

## Methods

### Study design and participants

This retrospective cohort study used the Twitter timelines—all of the tweets posted over time— of users who reported their pregnancy on Twitter. We identified these users via an automated natural language processing (NLP) tool, Pregex,^18^ that detects English-language tweets from the Twitter Streaming application programming interface (API) that self-report a gestational age or due date of an ongoing pregnancy and, based on the tweets’ timestamp, extracts dates marking the 40-week prenatal period. For example, the first tweet in Table 1 self-reported a gestational age of *20 weeks*, and, based on its timestamp, Pregex extracted the start date of pregnancy as August 13, 2021 and the due date as May 20, 2022. At the time each user was identified, we collected all of their available past tweets and began collecting all of their subsequent tweets on an ongoing basis. The timelines used in this study included tweets that were posted through August 2022. We re-deployed Pregex on each user’s full timeline in order to detect additional matching tweets, and used the tweets containing the most precise unit of time as the basis for further analysis.

**Table 1.** Sample tweets posted by a single Twitter user, illustrating self-reports of pregnancy (Tweet 1), COVID-19 infection (Tweet 2), preterm birth (Tweet 3), and maternal age (Tweet 4).

|  | **Tweet** | **Timestamp** |
| --- | --- | --- |
| 1 | @username @username Genuine question. I’m 20 weeks pregnant. Should I get the booster? Am I high risk or should I wait until I get my invite from the government? | 12-31-2021 |
| 2 | @username I mean, I got the vaccines, use the passport, wear a mask and sanitize my hands. I haven’t been in a large group in two weeks, and I work in an office with my own private space. But today, I tested positive for COVID. So what’s the point? | 03-02-2022 |
| 3 | @username My first had muscle weakness on one side of her mouth. I had to pump & bottle feed. I thought this baby would be different but shes a preemie and can’t latch. I’m pumping again & supplementing with formula. | 05-21-2022 |
| 4 | @username @username I’m 26 and already there. I just don’t care anymore. I’m going to do what I want and what makes me happy. | 02-20-2021 |

The sample tweets in Table 1 have been slightly modified to de-identify the user. The publicly available data used in this study were collected and analyzed in accordance with the Twitter Terms of Service. The institutional review boards of the University of Pennsylvania and Cedars-Sinai Medical Center reviewed this study and deemed it exempt.

### Exposures

The exposure for this study was COVID-19 infection during pregnancy. Therefore, we automatically excluded users with an extracted due date prior to March 11, 2020—the date that the World Health Organization (WHO) declared COVID-19 a pandemic. We deployed a deep neural network classifier to automatically detect tweets in the users’ timelines that self-reported a COVID-19 infection.^19^ The classifier was designed to identify only tweets that indicate an actual diagnosis, including a positive test, clinical diagnosis, or hospitalization. We manually validated the tweets that were automatically classified as a COVID-19 diagnosis, such as the second tweet in Table 1, excluding false positives. For true positives, we also manually validated the corresponding users’ tweets that were automatically detected by Pregex. We then used the timestamp and temporal textual features (e.g., *today, now, just, yesterday, on Friday, 3 weeks ago, on April 24*^*th*^, *at 36 weeks pregnant*) of the COVID-19 tweets to manually determine the timing of infection—first trimester (weeks 1-13), second trimester (weeks 14-27), or third trimester (weeks ≥28)—and exclude users with an infection that was not during pregnancy. For example, the timestamp and text of the second tweet in Table 1 indicate that the infection was during pregnancy—in particular, the third trimester. For tweets lacking explicit temporal features, we used the timestamp itself as an approximation of infection onset if we could infer from the text a temporal reference to the present. We excluded users with a COVID-19 infection that was during their pregnancy but for which we could not determine the specific timing.

### Outcomes

The outcome for this study was preterm birth. Therefore, we excluded users with a COVID-19 infection at ≥37 weeks of gestation—that is, beyond when preterm birth could occur. Among the manually validated users who were infected at <37 weeks of gestation, we deployed regular expressions—patterns of characters to search for matching text strings—to automatically detect tweets in their timelines that reported preterm birth,^20^ such as the third tweet in Table 1. For users who did not post matching tweets, we manually analyzed their timelines for other evidence of preterm birth, such as birth announcements with a timestamp or temporal textual features indicating that the baby was born more than three weeks prior to the due date. We excluded users who reported preterm birth prior to COVID-19 infection. Also, instead of assuming that the lack of tweets reporting preterm birth indicated that the pregnancy had reached term, we sought to mitigate this potential reporting bias by including term birth outcomes only for users with evidence of a gestational age ≥37 weeks. We automatically gathered some of this evidence by calculating the time difference between the due date and timestamp of the tweets that were detected by Pregex. For users who did not post a matching tweet within 21 days of their due date, we manually analyzed their timelines for other evidence of a gestational age ≥37 weeks.

### Covariates and control group

A covariate for this study that potentially could be identified on Twitter was maternal age. We deployed an automated NLP pipeline, ReportAGE,^21^ to extract users’ age from explicit self-reports in their tweets, such as the fourth tweet in Table 1. We manually validated the extracted ages and used the timestamp of the tweets to determine the users’ age at the time of their due date. For tweets that did not indicate the users’ birthday, we used a heuristic of rounding to the next year if the age was reported more than six months apart from the due date. For example, the user in Table 1 was aged 27 on February 20, 2022, which was less than six months away from a due date of May 20, 2022, so we estimated a maternal age of 27. To account for this covariate, we excluded users who did not post a tweet that was detected by ReportAGE, and we identified a 1:1 age-matched control group, consisting of users without COVID-19 infection during pregnancy. To select users for inclusion in the control group, we deployed ReportAGE on the timelines of users with an extracted due date prior to January 1, 2020. We manually validated the age matches among users with more recent due dates. We followed the same procedure for identifying outcomes in their timelines as we did for those who reported COVID-19 infection, excluding users without evidence of preterm birth or term birth. We re-sampled age-matched users until we identified an outcome for each one.

### Statistical analysis

We calculated the odds ratios (ORs) with 95% confidence intervals (CIs) to compare the overall rates of preterm birth for pregnancies with and without COVID-19 infection. We also performed subgroup analysis by comparing the control group with the rates of preterm birth by timing of infection: first trimester (weeks 1-13), second trimester (weeks 14-27), or third trimester (weeks 28-36).

## Results

Through August 2022, we automatically identified 153,038 users who self-reported their pregnancy on Twitter, and collected 1.1 billion tweets posted by these users. For 67,671 of these users, we automatically extracted a due date that was during the COVID-19 pandemic (Figure 1). Searching the timelines of these 67,671 users, we automatically detected 2075 tweets (1728 users) that self-reported a COVID-19 diagnosis. Through manual validation, we identified 501 users who were infected at <37 weeks of gestation. We identified a report of preterm birth for 38 (7.6%) of these 501 users, but excluded two of them who reported preterm birth prior to COVID-19 infection. Among the other 465 users, we identified 336 (72.3%) of them with evidence of term birth. Searching the timelines of the 372 users with evidence of preterm birth or term birth, we automatically detected and manually validated an age for 298 (80.1%) of them. The median maternal age for these 298 users was 26 years. For our age-matched control group, we searched for outcomes in the timelines of 363 users with a due date prior to January 1, 2020 in order to identify 298 users with evidence of preterm birth or term birth.

**Figure 1.**
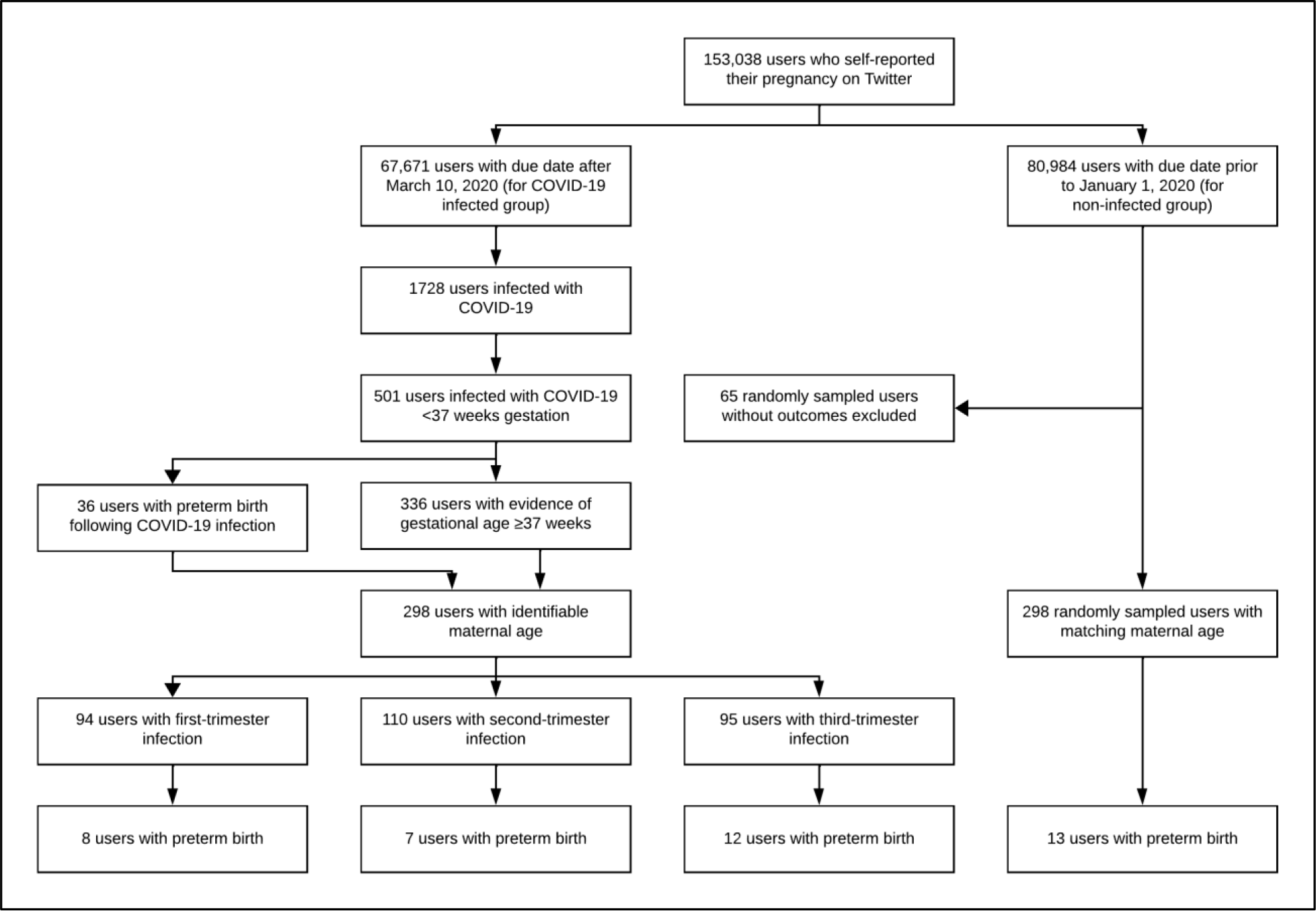
Study population selection.

Among the 298 users who met our inclusion criteria for COVID-19 infection, outcomes, and age, 268 (89.9%) of them reported a positive test, 21 (7%) of them reported a hospitalization, and 9 (3%) of them reported a diagnosis. Among these 298 users, 94 (31.5%) of them were infected during the first trimester (weeks 1-13), 110 (36.9%) of them were infected during the second trimester (weeks 14-27), and 95 (31.9%) of them were infected during the third trimester (weeks 28-36), with one user reporting separate infections in both the first and second trimesters. In total, 26 (8.8%) of these 298 users reported preterm birth: 8 (8.5%) who were infected during the first trimester, 7 (6.4%) who were infected during the second trimester, and 12 (12.6%) who were infected during the third trimester. In the control group, 13 (4.4%) of the 298 users reported preterm birth. In general, the risk of preterm birth was significantly higher for pregnancies with COVID-19 infection compared to those without (OR 2.1, 95% CI 1.06-4.16). In particular, the risk of preterm birth was significantly higher for pregnancies with COVID-19 infection during the third trimester (OR 3.17, CI 1.39-7.21). The risk of preterm birth was not significantly higher for pregnancies with COVID-19 infection during the first trimester (OR 2.04, 95% CI 0.82-5.08) or second trimester (OR 1.49, 95% CI 0.58-3.84) compared to those without infection.

## Discussion

The results of our study support that, in general, COVID-19 infection during pregnancy is associated with an increased risk of preterm birth. In particular, our results suggest that COVID-19 infection during the third trimester is associated with an increased risk of preterm birth. COVID-19 infection during the first or second trimester, however, did not appear to confer the same increased risk. The observed association between COVID-19 infection during pregnancy and preterm birth is consistent with recent systematic reviews and meta-analyses.^3–13^ Our results suggest, however, that the risk of preterm birth observed in prior studies may be driven by the preponderance of data on third-trimester COVID-19 infection^4,10,12^ and may not be generalized to infection during the first and second trimesters. To the best of our knowledge, our study included one of the largest samples of first-trimester COVID-19 infection. Even among the 2352 pregnancies with COVID-19 infection in Metz et al.’s study^22^ of one of the largest cohorts, only 54 (2%) of the infections were during the first trimester—approximately half the number of those in our study (n=94). Similarly, among the 1942 pregnancies with COVID-19 infection in Smith et al.’s meta-analysis^10^ of 12 studies, only 38 (2%) of the infections were known to be during the first trimester—less than half the number of those in our study. While the total number of COVID-19 infections in our study was coincidentally the same as that in Seif et al.’s study,^23^ the infections in our study were more evenly distributed across the trimesters and included nearly twice as many during the first trimester as theirs. Furthermore, along with Seif et al.’s study, our study was one of very few to assess the association of first-trimester COVID-19 infection with preterm birth.

In one of the few other studies that assessed the association between trimester of COVID-19 infection and preterm birth, Fallach et al.^24^ also found that pregnancies with third-trimester infection were at a higher risk of preterm birth, whereas pregnancies with first-trimester or second-trimester infection did not appear to be. In addition to supporting this finding, our study also complements theirs by addressing some of their noted limitations. As a limitation of their study, Fallach et al. wrote that, in their non-infected group, “women without positive SARS-CoV-2 test results did not necessarily test negative.” Therefore, it is possible that COVID-19 infections without test results were included in the non-infected group. In contrast, our control group ensured comparison with a non-infected group because it consisted of pregnancies with a due date prior to the COVID-19 pandemic. Because their study population was limited to Israel, Fallach et al. also noted that “[their] findings may have limited generalizability to countries populated with several races since there is evidence of disproportionate burden of COVID-19-related outcomes among different races.” While our study did not explicitly account for race, our prior work^15^ demonstrated that there are diverse races/ethnicities represented in our cohort of users who reported their pregnancy on Twitter, including White, Black, Asian, and Hispanic.

In contrast, Seif et al.^23^ found that COVID-19 infection during the first trimester was more likely to result in preterm birth than infection during the second or third trimester. They suggested that one possible explanation of their contrary findings could be related to the earlier time period of Fallach et al.’s study^24^: “Also, their study was conducted before July 2021 and, therefore, before the emergence of Delta and Omicron variants. Furthermore, due to the limited understanding of the disease process and how to manage it at the time of Fallach’s study, clinicians may have been more inclined to deliver COVID-19-infected patients nearing the end of their pregnancy.” However, the time period of our study—March 2020 to August 2022—extended even beyond that of Seif et al.’s. Therefore, their other explanation is more plausible: “One explanation could be that the study by Fallach et al. included all pregnant patients with a positive SARS-CoV-2 test. In contrast, we only included those who recovered from COVID-19 at the delivery time.” More generally, Seif et al.’s findings may not be directly comparable because our and Fallach et al.’s studies used a non-infected group for comparison, while Seif et al.’s study compared the trimesters to one another.

Our study has several limitations related to the use of Twitter data. First, some users reported their gestational age or due date only in terms of months. While some of these users may have been referring to a precise measure of time, others may have been approximating. In the case of the latter, the dates extracted by Pregex may have led us to misclassify the trimester of COVID-19 infection for some of these users. Second, while our control group selection—pregnancies with a due date prior to the COVID-19 pandemic—ensures comparison with a non-infected group, it does not account for potential pandemic-related confounders. Matching users based on negative test results, though, would have been more problematic because of a potential reporting bias on Twitter—namely, that users may have also tested positive but either did not report it or our methods did not detect it. Third, the rate of preterm birth reported in our control group (4.4%) was substantially lower than that in the general population around that time (10.2%),^25^ which might lead one to argue that Twitter does not represent the general population. Given that nearly half of adults aged 18-29 years in the United State use Twitter, ^26^ we believe that this discrepancy is more likely to reflect an underreporting of preterm birth on Twitter that does not differentially affect the infected and control groups. Fourth, because some users posted tweets reporting their age but not their birthday, using a heuristic to determine their age at the time of their due date involved a one-year margin of error for these age matches. Finally, while we were able to identify users’ age, the unmediated, self-report nature of Twitter data limited our ability to robustly model additional variables, such as illness severity, socioeconomic status, underlying diseases, parity, COVID-19 vaccination status, and whether the preterm birth was medically indicated (i.e., infection was the reason for delivering preterm) or spontaneous.

In summary, we used large-scale, longitudinal social media data in a retrospective cohort study to assess the association between the trimester of COVID-19 infection and preterm birth. We found that COVID-19 infection particularly during the third trimester is associated with an increased risk of preterm birth.

## Data Availability

According to the Twitter Terms of Service, the content (e.g., text) of Tweet Objects cannot be made publicly available, and the number of tweets that were analyzed in this study exceeds the total that can be made publicly available as Tweet IDs; however, a limited number of Tweet Objects are permitted to be shared directly, and an unlimited number of Tweet IDs are permitted to be shared on behalf of an academic institution for non-commercial research. Requests for Tweet Objects or Tweet IDs can be sent to Ari Z. Klein or Graciela Gonzalez-Hernandez.

## Contributors

AZK and GGH conceptualized the study. AZK, JCF, and GGH designed the study. AZK collected the data. AZK and SK analyzed the data. AZK and LDL interpreted the results. SG performed the literature search and statistical analysis. AZK drafted the manuscript. All authors reviewed the manuscript.

## Acknowledgments

This work was supported by the National Library of Medicine (R01LM011176). The content is solely the responsibility of the authors and does not necessarily represent the official views of the National Institutes of Health. The authors thank Ivan Flores for contributing to software applications, and Kaelen Spiegel for contributing to data analysis.

